# A SYSTEMATIC REVIEW AND META-ANALYSIS ON THE EFFECT OF CURCUMIN (TURMERIC) ON FASTING BLOOD GLUCOSE AND GLYCATED HAEMOGLOBIN IN PATIENTS WITH TYPE 2 DIABETES MELLITUS

**DOI:** 10.1101/2023.10.13.23297012

**Authors:** David Chinaecherem Innocent, Rejoicing Chijindum Innocent, Ramesh Kumar, Ali A. Rabaan, Chiagoziem Ogazirilem Emerole, Oluwaseunayo Deborah Ayando, Ihuoma Chimdimma Dike, Chinazaekpere Oguguo Duruji

## Abstract

**Background:** Hyperglycemia is a recurring metabolic condition known as diabetes mellitus. When glycated haemoglobin (HbA1c) >6.5% and fasting blood glucose (FBG) continuously falls below 126 mg/dl, it is clinically diagnosed. Type 2 diabetes (T2DM) and the emergence of diabetic complications are mostly mediated by oxidative stress and chronic inflammation. Research on the use of natural anti-inflammatory and antioxidant substances, such as curcumin (turmeric), as an adjuvant treatment in the management of T2DM is becoming more and more popular. However, the effects of curcumin on glycemic control in T2DM patients have varied according to the outcomes of randomised control trials. Therefore, this systematic review/meta-analysis was aimed at synthesizing findings from different RCTs to determine the effect of curcumin on fasting blood glucose and glycated haemoglobin in patients with T2DM.

**Method:** Searches were conducted in electronic databases and other sources such as PubMed, CINAHL, CENTRAL, ProQuest, Web of Science, Health Technology Assessment (HTA), Scopus, LILACS, clinicaltrials.gov and google scholar. Key search terms use included curcumin, fasting blood glucose, glycated haemoglobin, and type 2 diabetes mellitus. Relevant RCTs conducted within the last 12 years (2009-2022) were selected and assessed against the inclusion criteria. A summary of the search strategy was presented on a PRISMA flow chart. Data were extracted using standardised data extraction forms and meta-analysis was performed using the RevMan version 5.3. and results presented using forest plots.

**Results:** Five RCTs with a total of 349 participants were included in the meta-analysis. Curcumin supplementation significantly reduced fasting blood glucose and glycated haemoglobin in patients with type 2 diabetes mellitus when compared with the usual drugs. FBG (MD: -1.84, 95% CI: -4.92 to 1.24, P=0.24, I^2^=15%). HbA1c (MD: -0.24, 95% CI: -0.55 to 0.07, P=0.13, I^2^=0%).

**Conclusion:** Curcumin supplementation is effective in the management of T2DM and diabetic complication. Further research on ways to bypass the challenges of bioavailability such as the use of nano-micelles may produce greater therapeutic effects on diabetes management.

## 1.0 Introduction

Hyperglycemia is a recurring metabolic disease known as diabetes^1^. Clinical diagnosis is made when glycated haemoglobin (HbA1c) is consistently 48mmol/mol or >6.5% and fasting blood glucose is consistently 126mg/dl ^2^. According to the World Health Organization (WHO), 2019 polyphagia, polyuria, and polydipsia are the three main signs of diabetes^3^. Endothelial dysfunction brought on by long-term type 2 diabetes mellitus (T2DM) results in cardiovascular illnesses and tissue abnormalities such retinopathy and nephropathy. This has significantly aided in the rise in diabetes-related morbidity and death worldwide^4^. Obesity/overweight is the most important predisposing factor to T2DM. Also, over 600 million people are reported to be affected by obesity and 90% of diabetic patients are reported to be overweight^5^. Obesity accounts for 60-80% of type 2 diabetic cases in Africa and Europe^6^.

Diabetes is currently ranked as the 7^th^ leading cause of death globally accounting for about 1.6 million deaths per annum^7^. It is also reported that over 422 million people have diabetes globally with higher incidence occurring in low to middle-income countries. Type 2 diabetes mellitus make up 90% of all cases of diabetes worldwide and hyperglycaemia causes about 4 million death annually^8^. Estimation by WHO suggest that these figures are expected to double by 2040 thus, the intensified global effort to stop the rise of diabetes and obesity by 2025^9^. Africa and Asia are predicted to have the highest incidence by 2030 largely due to changes in lifestyle, late detection and socio-economic factors^10^. Over 60% of diabetic patients are reported to be from Asia however, cases are recorded in virtually all countries of the world^7,11^. It is also estimated that 30%-80% of diabetic cases are either are diagnosed lately or not diagnosed at all.

Diabetes being a major public health concern also poses a huge economic burden on individuals and society. The average expenditure per diabetic patient is estimated at $1583 largely due to the expensive branded/patented medications^12^. In 2017, the global economic cost of diabetes was $850 billion, yet it is reported that this money falls short of the money needed to reach the global target by 2025^13^. Diabetic complications also create an economically inactive population further leaving adverse socio-economic effects on the patients and their dependants especially in developing countries^14^. These huge health and economic burden have called for concerted efforts and intensified research into cheaper and more effective interventions for the management of diabetes and diabetic complications^15^.

The current interventions for T2DM usually involve the prolonged use of oral anti-hyperglycaemic agents such as metformin, injectable glucagon-like peptide-1 (GLP-1) receptor agonists e.g. exenatide, and non-chemical interventions such as diet modification and increased physical activities^16^. These common anti-hyperglycemic agents help to either maintain blood glucose levels, increase insulin sensitivity of cells or stimulate the pancreatic islet cells to produce more insulin^17^. They are said to prolong and increase the quality of life of patients with T2DM^18^.

However, there are concerns as to their cost-effectiveness and the numerous adverse effects (e.g. pancreatitis and pancreatic cancer). Also, shreds of evidence show that these chemical drugs aid fat accumulation, causes hypoglycaemia, hypovolaemic shock and other adverse effects associated with prolonged usage^19,20^. These drugs have little or no effect on oxidative and inflammatory cellular processes that are largely linked with the development of hyperglycaemia, diabetes and diabetic complications^21,22,23^. This has called for safer, cheaper and more potent alternatives or supportive interventions such as antioxidant compounds of natural plant origin for the management of diabetes^24^.

Given the advancement in medical science and technology, interventions such as bariatric surgery (commonly used for obese diabetic patients), pancreatic transplant and digital health have demonstrated patient’s improvement and increased adherence to treatment^25,26^. However, transplantation and bariatric surgery come with an increased risk of complications and they are not cost-effective^27,28,29^.

In recent years, there has been increased attention to research on alternative interventions for T2DM. One of which include emerging research on the effects of antioxidants as adjunct interventions in the management of hyperglycaemia and diabetes^30^.Ceriello suggested that antioxidants may be the supportive intervention of interest to help fix the underlying cause of T2DM and diabetic complications^31^.

Various research has demonstrated that hyperglycaemia is largely linked to chronic inflammation and oxidative stress processes which results in the release of highly reactive molecules such as reactive oxygen species (ROS) and reactive nitrogen species (RNS) ^22,32^. Although the roles of ROS and RNS in T2DM have not been fully understood, studies show that these free radicals and the inflammatory processes impair the insulin signalling pathway which leads to insulin resistance or poor insulin sensitivity hence, the development of chronic hyperglycaemia and T2DM^33^. Studies have also shown that diabetic patients have a significantly reduced antioxidants levels and higher levels of free radicals but the common interventions are not able to correct or mitigate these pathologies^34,35^.

Natural antioxidants and anti-inflammatory compounds such as turmeric have been used for centuries in the treatment of different diseases in traditional medicine^36^. However, there is a paucity of scientific evidence on their usage in the management of chronic diseases such as T2DM^37^. With more studies carried out on animal studies, only a few pieces of evidence are available to prove the effectiveness of natural antioxidants in the treatment of T2DM in studies with human participants^38^. More so, several mixed evidence exists from the few human studies posing a huge challenge in clinical decision and health policy formulation concerning diabetic management. These unanswered questions and gap in knowledge have prompted an interest in conducting this review.

Fasting blood glucose (FBG) is a measure of blood glucose concentration. It gives information about the physiologic integrity of insulin-signalling pathways, glucose metabolism and viability of insulin-associated tissues^39^. Usually, patients with FBG ≥ 126mg/dl or 7mmol/l are said to be hyperglycaemic and considered to have diabetes if consistent^2^.

On the other hand, HbA1c gives an indication of the percentage of blood glucose attached to haemoglobin. It gives the average blood sugar level within a 2-3 months period expressed in percentage^40^. A percentage of ≥6.5% indicates that the person has diabetes and higher percentages show the progression of the pathology and higher risk of developing diabetic complications^2^. In hyperglycaemic patients, more haemoglobin binds to blood sugar which results in poor tissue oxidation, production of free radicals and damage to insulin-producing cells. Also, endothelial pathologies usually lead to tissue damage and organ failures^40^.

Fasting blood glucose and HbA1c are clinically the two most important diagnostic parameters for diabetes^41^. They are also critical in monitoring the progression of diabetes as well as determining the effect of any diabetic intervention, which informs their measurement in many interventional studies regarding diabetes and glycaemic control. Therefore, conducting a systematic review and meta-analyses of RCTs that measure these parameters has become imperative in providing substantial evidence on the therapeutic roles of curcumin (turmeric) on glycaemic control and management of T2DM.

RCTs and systematic reviews are topmost on the hierarchy of evidence. Systematic reviews pool evidence from various studies into a few sources that can easily be accessed to provide information on the effectiveness of the intervention and inform decision making^42^. Therefore, this systematic review and meta-analyses are aimed at increasing the strength of evidence on the effect of curcumin as an adjunctive intervention in the management of hyperglycaemia and T2DM. This was carried out by pooling pieces of evidence from various randomised control trials conducted within a 10-year period (2009-2019). A meta-analysis is then conducted and results of the effect estimates are presented on a forest plot.

### Research question

To ensure this question is properly answered, the standardized PICO (Population, Intervention, Comparator and Outcome) framework was used to define the research question thus, the review question is: Does curcumin have any effect on fasting blood glucose and glycated haemoglobin in patients with T2DM?

### Aim and Objectives

While the aim of a study establishes the overall goal or expected outcomes, objectives of the study are the steps taken to accomplish the desired goal. The aim and objectives of this study were clearly stated using the research question as a guideline.

### Aim

To establish evidence on the effect of curcumin on fasting blood glucose and glycated haemoglobin in patients with T2DM.

### Objectives

□ To explore current and existing knowledge on the antioxidant and anti-inflammatory roles of curcumin and its therapeutic effect on diabetics, given that oxidative stress and chronic inflammation plays a critical role in the pathogenesis of T2DM.

□ To conduct a systematic review and meta-analysis on different RCTs carried out in the last 10 years (2009-2019) on the effect of curcumin on glycaemic control in patients with T2DM.

□ To increase the sample size thus, increasing the statistical power, strength and precision of evidence by pooling different RCTs conducted on the research question.

□ To also explore information on the therapeutic dose and duration in which curcumin is effective in the management of hyperglycaemia in patients with T2DM.

□ Lastly, using statistical software packages such as Revman to present results and the pooled effect estimate which will establish the effectiveness of curcumin as an intervention to manage T2DM.

## 2.0 Methods

In order to ensure standardization, quality, transparency and replicability of this review, the method employed must be clearly stated. The PRISMA checklist, the Cochrane andCASP guidelines for conducting a systematic review and meta-analyses to determine the effect of clinical interventions were followed in this review.

### Eligibility of studies

The eligibility in this study was based on certain characteristics that qualify the RCTs to be included in this systematic review and meta-analysis. As suggested by Liberati et al. (2009), this was determined by clearly setting out the inclusion and exclusion criteria which were largely guided by the PICO framework, the research question and the PRISMA checklist as stated below.

### Type of studies

Only RCTs conducted from 2009-2019 showing the effect of curcumin/turmeric on fasting blood glucose and glycated haemoglobin in patients with T2DM were included in the analyses. RCTs included those published in English language, unpublished pieces of literature and in non-English journals such as LILACS. This is done to ensure enough coverage of all accessible articles and minimize the possibility of biases.

### Participants

RCTs are those conducted on patients clinically diagnosed with T2DM onlyi.e. fasting blood glucose levels ≥126g/dl at the beginning of the study. Studies whoseparticipants have other disease conditions or comorbidities were excluded. Participants are adults ranging from age 18-75 years. Studies conducted on participants in critical care receiving other medications were not included. Also, similar studies on animal models were excluded.

### Intervention

Included studies were those in which curcumin is the only experimental intervention administered to the treatment group in addition to the usual drugs such as metformin.

### Comparator

The comparator in this study involves the administration of the usual oral hypoglycaemic agents such as metformin to the control group. This was conducted based on the ethical recommendations in line with the Helsinki declaration and research standards for diabetic patients. The aim is to compare the effect of curcumin supplementation against the usual diabetic drugs given to patients with T2DM.

### Outcomes

The primary outcomes of interest in this study were fasting blood glucose and glycated haemoglobin (HbA1c). These were measured because they are used to indicate the therapeutic effect of diabetic interventions in this case curcumin in patients with T2DM. RCTs which measure multiple outcomes but included fasting blood glucose and HbA1c major outcomes were also included provided the study meets the criteria for inclusion.

### Setting

Studies were not restricted to any geographical part of the world however, they must follow standard clinical trial procedures.

### Search strategy

A systematic and carefully conducted search strategy is necessary to pool out studiesthat will effectively answer the research question^43^.A pilot search was first conducted based on PICO and the research question to identify existing knowledge and to avoid repetition of studies. The search was carried out using an electronic database such as PubMed, CINHAL, Medline, the web of science, and clinicaltrials.gov. Search engines like Google scholars were used to extract references and to access full texts.

To limit bias and to widen the scope of the search, a non-English database such as LILACS was also searched. Unpublished works of literature were obtained from the Health Technology Assessment (HTA) database and grey literature such as international conference papers on diabetes. Manual searching of journals and the reference list of relevant articles were also carried out. The first search was conducted between June 1^st^,2019 and July 30^th^, 2019 using the different databases as shown in table 1 below.

Key search terms, Boolean operators, wildcards and truncations were used to search the different databases in order to expand the scope of literature search and identify the relevant literature. Examples of these include (“curcumin” OR “turmeric/tumeri* OR “polyphenol” OR “antioxidants”) AND (“glycated haemoglobin and blood glucose concentration” OR”blood sugar levels” OR “fasting blood glucose” OR “hyperglyc#emia”) AND (“T2DM” OR “diabetes” OR “hyperglycaemia” OR “insulin-resistant diabetes”).

### Study selection

To reduce the risk of bias, it is recommended that two independent researchers carry out the selection process. However, as a fulfilment for the master’s programme, only one reviewer conducted the process under the guidance of a more experienced supervisor.

In the first stage of the selection process, the titles and abstracts of the searched pieces of literature were screened to identify the most relevant studies from the various databases. A referencing software (endnote) was used to collate, organize and remove duplicated studies. However, due to minor differences in indexing references on the different databases, a manual approach was employed to identify and removed duplicates that were missed out in by endnotes as suggested by^41^.

At the second stage, full texts of relevant studies were carefully analysed against the inclusion and exclusion criteria for eligibility. Studies which were not eligible were excluded from the meta-analyses with reasons stated. A summary of the selection process is presented on a PRISMA flowchart in figure 1 below.

**Figure 1.0:**
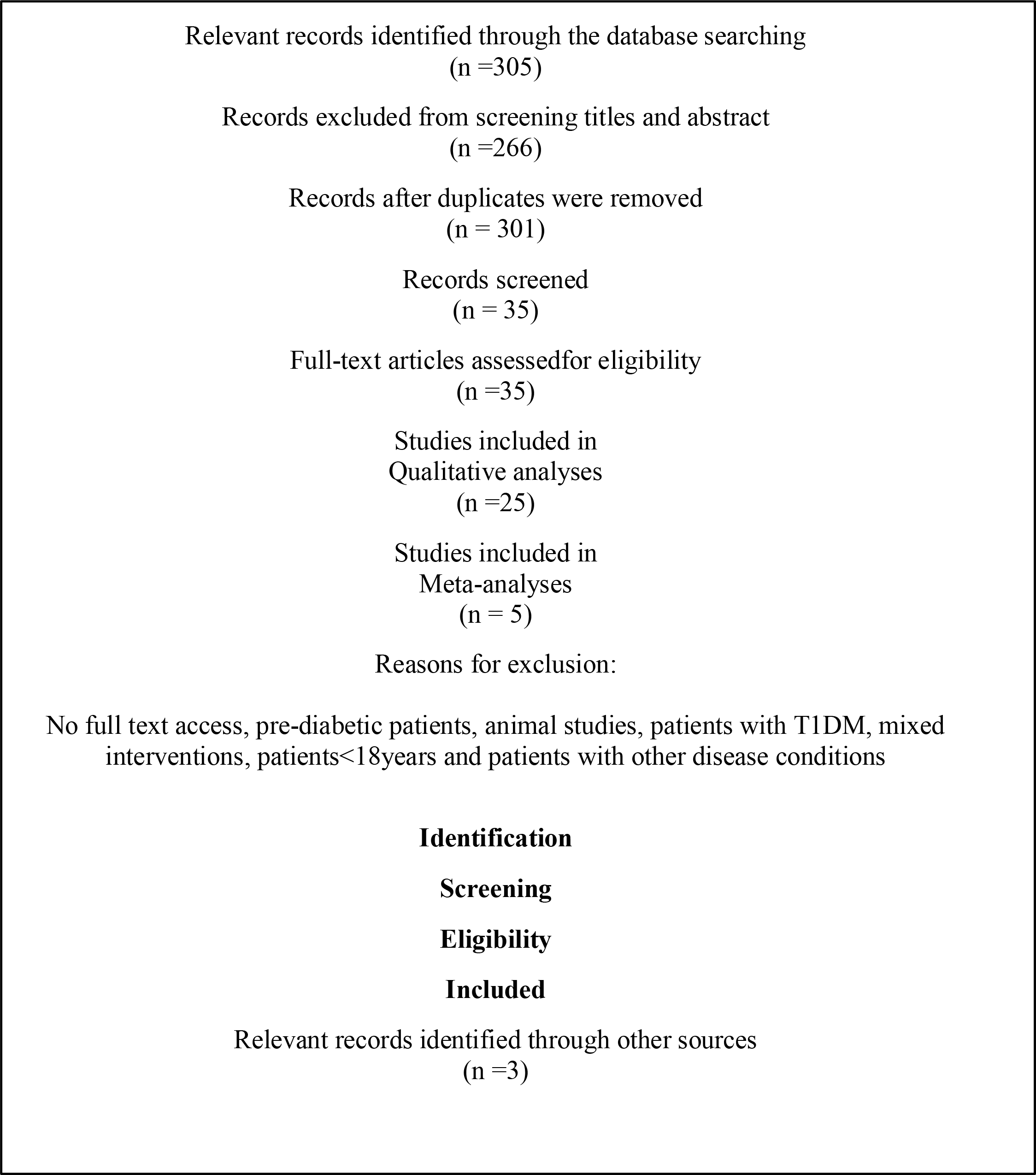
PRISMA flow chart

**Figure 2.0:**
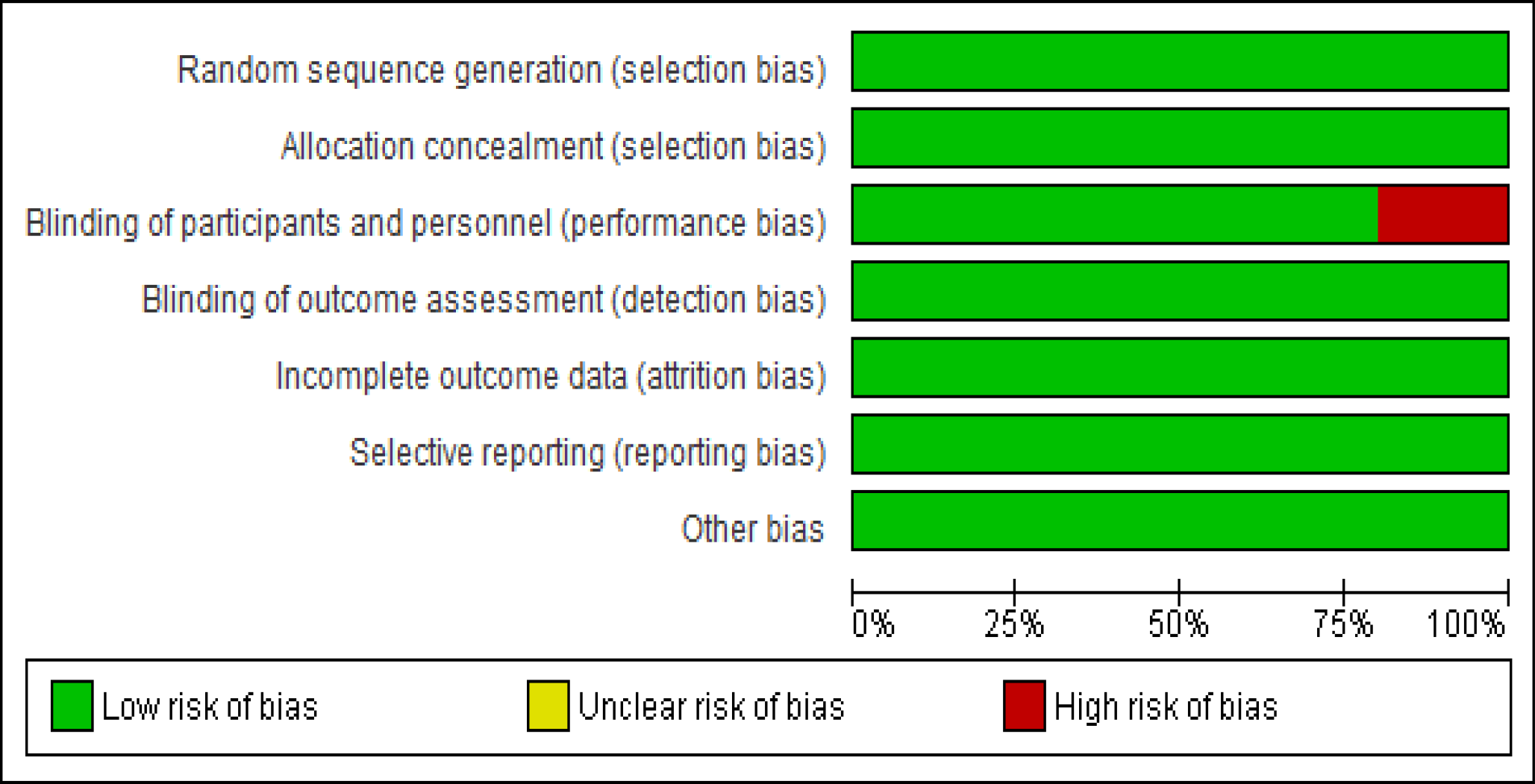
Risk of bias graph

**Figure 3.0:**
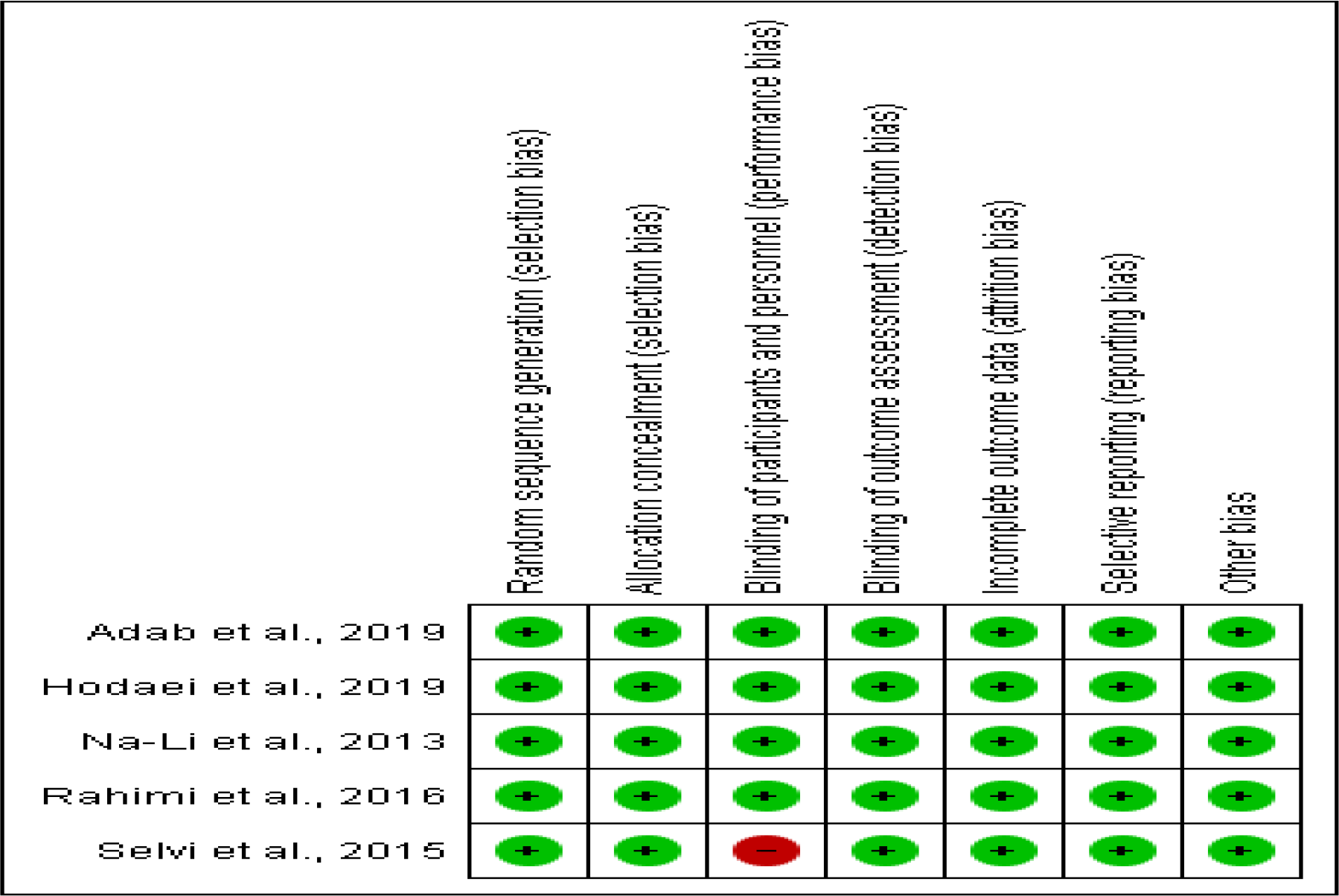
Risk of a bias graph summary^47,44,48,46,43^

### Quality Assessment

Although RCTs are the gold standard for measuring the effects of the interventionand not all RCTs are classed as high-quality studies which can be used to conduct a systematic review and meta-analyses. Therefore, a quality assessment is critical to ensure substantial, quality and generalizable evidence^41,42^. There are different quality assessment tools used to assess the validity of a RCTs research paper such as the Jadad scale, Cochrane collaboration and the CASP tool. A higher quality of RCTs will give better credibility to the outcomes of the metaanalysis. In the current study, the Jadad scale and CASP tool were used to assess the quality of the included studies as shown in table 2 and table 3 below respectively. From the Jadad score, all except one of the included studies demonstrated high-quality studies represented by a Jada score of 5. The study by Selvi et al. (2015) demonstrated a lower Jadad score of 3. This is because the study was a single-blinded RCT, however, the paper is still considered to have a good quality for inclusion in this review^43^.

**Table 2.0:** Jadad score for quality assessment of included studies^47,44,48,46,43^

| STUDY/YEAR | RANDOMISATION |  | BLINDING |  | FOLLOW UP | TOTAL SCORE |
| --- | --- | --- | --- | --- | --- | --- |
|  | Study randomised? | Method for random sequence generation described and appropriate? | Described as double-blind? | Method of double-blinding described and appropriate? | Withdrawal/dropout described? |  |
| Adab et al. (2019) | 1 | 1 | 1 | 1 | 1 | 5 |
| Hodaei et al. (2019) | 1 | 1 | 1 | 1 | 1 | 5 |
| Na-Li et al. (2013) | 1 | 1 | 1 | 1 | 1 | 5 |
| **Rahimi et al. (2016)** | 1 | 1 | 1 | 1 | 1 | 5 |
| **Selvi et al. (2015)** | 1 | 1 | 0 | 0 | 1 | 3 |

**Keys:** 1 = pass, 0 = fail

**Table 3.0:** CASP tool for quality assessment of eligible studies^47,44,45,46,43^

| SN | STUDY | 1 | 2 | 3 | 4 | 5 | 6 | 7 | 8 | 9 | 10 | 11 |
| --- | --- | --- | --- | --- | --- | --- | --- | --- | --- | --- | --- | --- |
| 1 | **Adab et al. (2019)** | Y | Y | Y | Y | Y | Y | FBG and HbA1C levels are reduced but not statistically significant. | FBG p=0.57<br>HbA1c p=0.90 | Y | Y | Y |
| 2 | **Hodaei et al. (2019)** | Y | Y | Y | Y | Y | C | FBG significantly decreased<br>HbA1C levels no significant decrease | FBG p=0.05<br>HbA1c p=0.3 | Y | Y | Y |
| 3 | **Li-Xin et al. (2013)** | Y | Y | Y | Y | Y | C | FBG significant decrease<br>HbA1C reduced but not statistically significant | FBG p<0.01<br>HbA1c p=0.031 | Y | Y | Y |
| 4 | **Rahimi et al., 2016** | Y | Y | Y | Y | Y | C | FBG and HbA1C levels are significantly reduced | FBG p=0.004<br>HbA1c p<0.001 | Y | Y | Y |
| 5 | **Selvi et al. (2015)** | Y | Y | Y | Y | Y | Y | FBG and HbA1C levels are significantly reduced | FBG p<0.001<br>HbA1c p=0.044 | Y | Y | Y |

**KEYS:** Y =Yes, N =No, C = Cannot tell

**Table 4.0:** Summary of extracted data^47,**44**,**48**,**46**,**43**^

| Study/<br>Year | Sample size | Loss to follow-up | Duration of study | Study type | Age of participants | Health status | Setting | Country of study | Intervention/dose | Outcome measures |
| --- | --- | --- | --- | --- | --- | --- | --- | --- | --- | --- |
| Adab et al. (2019) | 80 | 5 | 8 weeks | Double-blind RCT | 30-70 years | T2DM patients | Metabolism Research Centre in Firoozgar Hospital of Iran University of Medical Sciences | Iran | Turmeric 700mg thrice daily (total of 2100mg) | FBG and HbA1c |
| Hodaei et al. (2019) | 53 | 9 | 10 weeks | Double-blind RCT | 40-70 years | T2DM patients | National Nutrition and Food Technology Research Institute of Shahid Beheshti University. | Iran | Curcumin 500mg thrice daily (total of 1500mg) | FBG and HbA1c |
| Na-Li et al. (2013) | 109 | 9 | 12 weeks | Double-blind RCT | 18- 65 years | T2DM patients | 2nd Affiliated Hospital of Harbin Medical University, China in 2009. | China | Curcuminoids 150mg twice daily (total of 300mg) | FBG and HbA1c |
| Rahimi et al. (2016) | 80 | 10 | 12 weeks | Double-blind RCT | $\geq 18$ years | T2DM patients | Mashhad University of Medical Sciences. | Iran | Nano-curcumin 80mg once daily | FBG and HbA1c |
| Selvi et al. (2015) | 60 | 0 | 4 weeks | Open-label RCT | 35-55 years | T2DM patients | Jawaharlal Institute of Postgraduate Medical Education and Research | India | Turmeric 2000mg once daily | FBG and HbA1c |

**Table 5.0:** Data extracted on the effect of curcumin on FBG^47,44,48,46,43^

| Study ID/ Year | Mean ± SD in the curcumin group | Sample size analysed in the curcumin group | Mean ± SD in the control group | Sample size analysed in the control group | Total sample size completely analysed |
| --- | --- | --- | --- | --- | --- |
| Adab et al. (2019) | 131.64 ± 28.33 | 39 | 139.41 ± 41.57 | 36 | 75 |
| Hodaei et al. (2019) | 153 ± 33 | 21 | 147 ± 40.4 | 23 | 44 |
| Na-Li et al. (2013) | 7.28 ± 1.77 | 50 | 8.17 ± 2.06 | 50 | 100 |
| Rahimi et al. (2016) | 120.29 ± 38.01 | 35 | 176.0 ± 61.56 | 35 | 70 |
| Selvi et al. (2015) | 95 ± 11.4 | 30 | 102 ± 18 | 30 | 60 |

**Table 6.0:** Data extracted on the effect of curcumin on HbA1c^47,44,45,46,43^

| Study ID/ Year | Mean ± SD in the curcumin group | Sample size analysed in the curcumin group | Mean ± SD in the control group | Sample size analysed in the control group | Total sample size analysed |
| --- | --- | --- | --- | --- | --- |
| Adab et al. (2019) | 7.04 ± 0.98 | 39 | 7.28 ± 1.59 | 36 | 75 |
| Hodaei et al. (2019) | 11 ± 2 | 21 | 11.1 ± 1.8 | 23 | 44 |
| Li-Xin et al. (2013) | 7.02 ± 2.04 | 50 | 7.99 ± 2.86 | 50 | 100 |
| Rahimi et al. (2016) | 7.31 ± 1.54 | 35 | 9.00 ± 2.33 | 35 | 70 |
| Selvi et al. (2015) | 7.4 ± 0.9 | 30 | 7.5 ± 0.7 | 30 | 60 |

From the CASP checklist, all 5 included studies showed good quality for a systematic review. However, studies by Hodaei et al. (2019), Li-Xin et al. (2013) and Rahimi et al. (2016) were not detailed on which conventional medication was given to the control group^44,45,46^.

### Risk of Bias Assessment

Conclusions on the effect of interventions in clinical trials and systematic review are dependent on the research design and validity of included studies respectively. Validity is a crucial consideration in systematic review which influences the effectiveness, generalisability of results and clinical decision on interventions. However, bias poses a great threat to the validity of the study outcome in clinical trials and SRs thus, the need to assess for risk of biases. Bias is usually a systemic or methodological error that can overestimate or underestimate the true effect of an intervention.

In the current study, the classification of bias by Cochrane collaboration was used to assess the risk of bias of the included studies. These include selection bias, performance bias, detection bias, attrition bias and reporting bias.

Selection bias was assessed by analysing for the method of sequence generation and allocation concealment. All five studies reported the allocation concealment and randomisation process. Performance bias was assessed by checking for blinding. Four of the studies reported the blinding of participants. The study by Selvi et al. was an open-label study which gives a performance bias^48^. Detection bias was assessed by analysing for blinding of outcome concealment. It was unclear whether the study by blinded participants to the outcome assessors. Incomplete outcome data was verified to reduce the risk of attrition bias. For instance, the study by Na et al. did not state the exact hypoglycaemic treatment received by the participants in the curcumin group^46^. Lastly, all the studies were also analysed for reporting bias by looking out for selective outcome reporting.

### Data extraction and analysis

Data extraction in a systematic review is the process of analysing and retrieving and analysing pertinent data from primary quantitative research using a standardized data extraction form or table. In this study, RevMan used in Cochrane reviews was adopted. One reviewer extracted the data which included information about the study characteristics, the subject and outcome measure.

Data extracted showing the effect of interventions in all studies were given as mean ± standard deviation of endpoint values in the intervention and control groups. Where studies have multiple outcomes, only treatment effects on the outcome of interest (FBG and HbA1c) were extracted for analyses. Variations in the units of measurement were reconciled by conversion to the same units. The treatment effect displayed as mean ± SD were rounded up to one decimal place before inputting into RevMan to give uniformity. The sample size used in RevMan only included participants that completed the studies and were analysed in both the experimental and control group.

Since data on outcomes are continuous (i.e. a measure of HbA1c and FBG concentration), a continuous data model and summary statistics (effect measure) of the mean difference were selected in the RevMan software to estimate the pooled effect and presented on the forest plot (see figure 4 and 6 below). The analysis was carried out using the random effect model. Data were displayed as the weighted mean difference with a 95% confidence interval (95%CI). The random effect analysis tool adopted was the inverse variance. Also,the result on effect size was presented on funnel plots for both fasting blood glucose and HbA1c (see figures 5 and 7 respectively).

**Figure 4.0:**
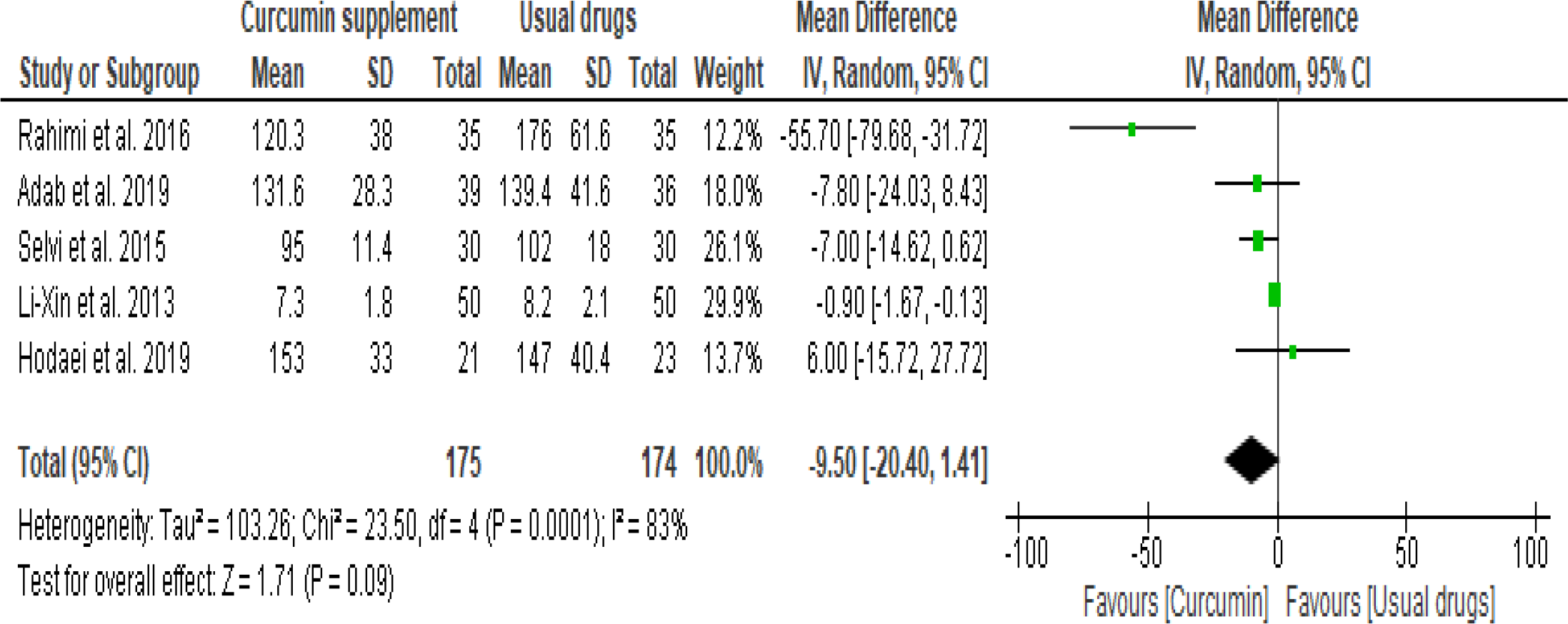
Forest plot of comparison on FBG before sensitivity analysis^46,47,43,45,44^

**Figure 5.0:**
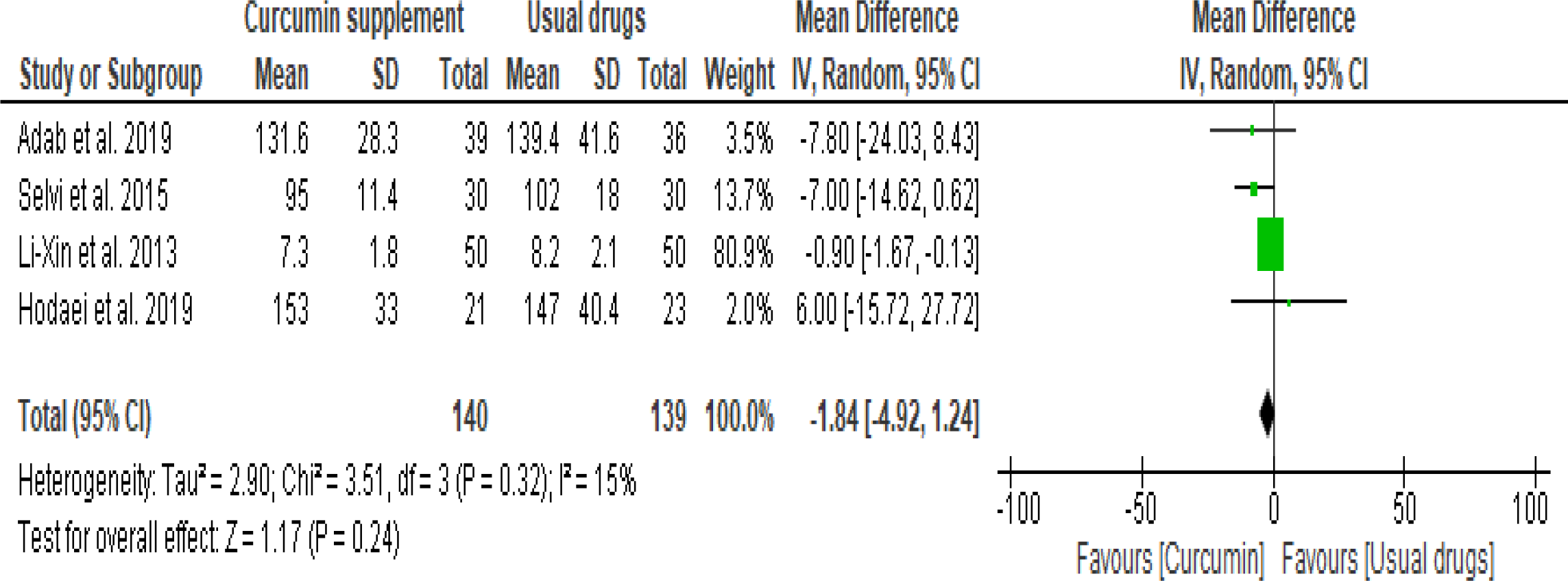
Forest plot of comparison on FBD after sensitivity analysis^47,43,45,44^

**Figure 6.0:**
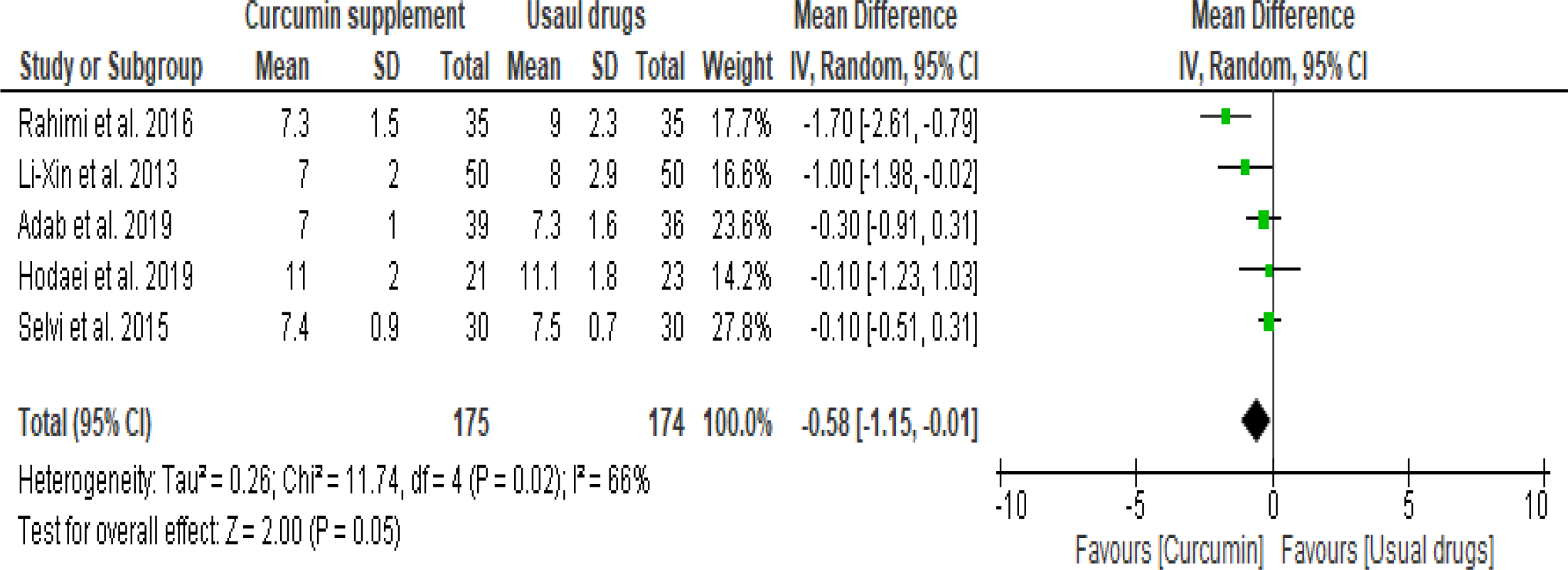
Forest plot of comparison on HbA1c before sensitivity analysis^46,45,47,44,43^

**Figure 7.0:**
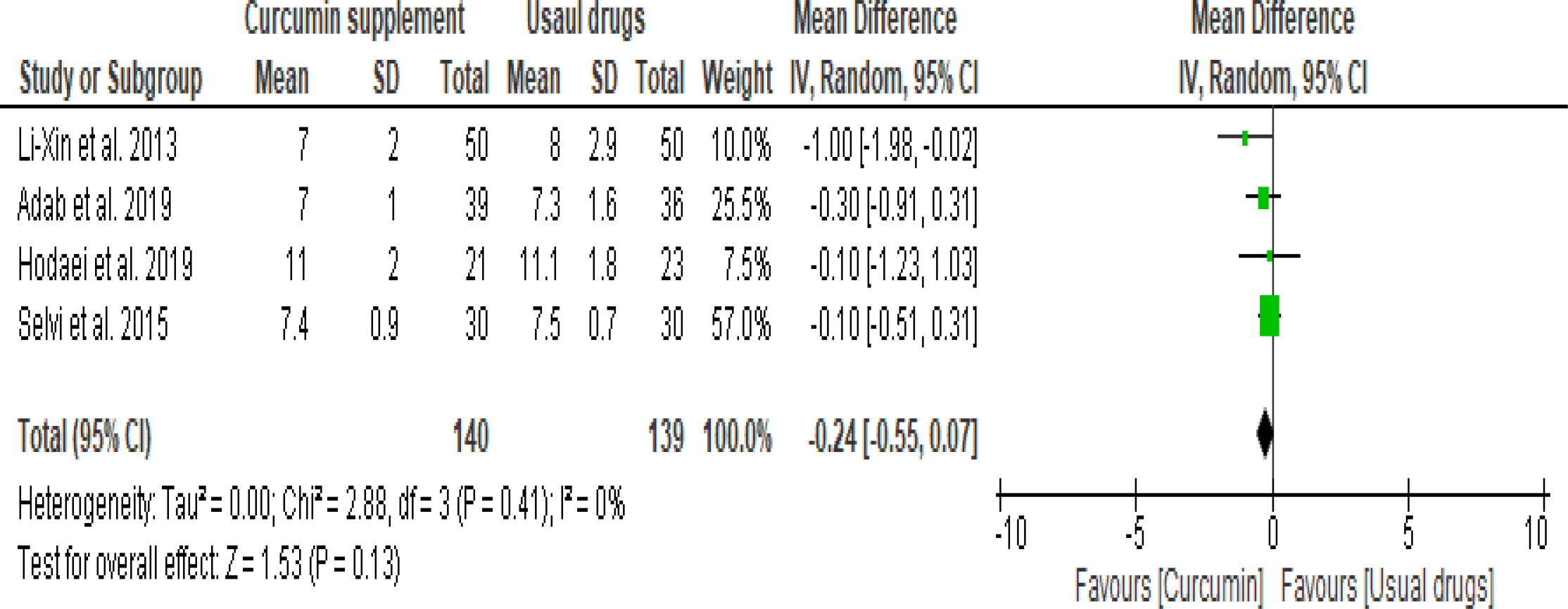
Forest plot of comparison on HbA1c after sensitivity analysis^45,47,44,43^

I-squared statistics (I^2^) which measures the degree of variation in the different studies were used to test for heterogeneity. A significant degree of heterogeneity was represented by I^2^>50% and p-value <0.1. Also, any p-values ≥ 0.05 on the overall results or subgroup results were considered to have no statistical significance.

## 3.0 Results

Following the search from electronic databases, 305 relevant records were identified. Further search from other sources resulted in 3 additional studies bringing the total number of studies to 308. 7 duplicates were removed using endnote. One of the additional studies was a non-English paper which was translated into the English language. The titles and abstracts of 301 studies were screened out of which the full texts of 35 studies were assessed for eligibility and inclusion.

Of the 35 studies assessed, 25 studies were excluded from the meta-analyses but included in the qualitative analyses because of the following reasons: Non-full text access, pre-diabetic patients, animal studies, patients with T1DM, mixed interventions, patients<18years, patients with other disease conditions and non-RCT studies. Finally, only 5 studies qualified and were included for meta-analyses.

### Results from meta-analyses

On the overall, five RCTs were used in the meta-analyses with a total of 349 participants (175 participants in the treatment/curcumin group and 174 participants in the control group). Following the extraction and analyses of the inputted data from the Revman 5.3 statistical software, the results were presented on forest plots and funnel plots as shown above.

### Results from forest plots

From the forest plotsabove, thehorizontal lines to the right represent the confidence intervals for each study. The longer the line, the wider the confidence intervals and the less precise the result. The boxes on each horizontal line represent the point estimates for each study. The sizes of the boxes indicate the weight contributed by each of the included study, the wider the box, the greater the weight contributed. The diamond figure represents the pooled or overall effect estimate of all included studies. The right and left wings of the diamond indicate the confidence intervals while the top and bottom points indicate the overall point estimate. The wider the diamond, the wider the confidence interval and the less precise the result.

Also, the vertical line on point zero is the line of no effect or line of no significance. Individual studies whose point estimates falls towards the left of the line of no effect favours the curcumin supplemented group while studies that fall towards the right favours the control group where the usual drugs were administered. At the bottom of the tables on the forest plots are information on the heterogeneity (Chi^2^ and I^2^), and test of overall effect (Z). A significant degree of heterogeneity was represented by I^2^>50% and Chi^2^ with a p-value <0.1 while the reverse shows little or no variability across the included studies. Any overall effect with p<0.05 is considered to be statistically significant.

The first forest plot in figure 4.0 below shows results for the effect of curcumin on fasting blood glucose. The point estimate for all the studies falls to the left of the line of no effect in favour of curcumin, except the study by Hodaei et al. 2019 which favoured the usual drugs^44^. The overall effect estimate represented by the diamond falls to the left of the line of no effect in favour of patients supplemented with curcumin having an overall mean difference/95% confidence interval of -9.50 (-20.40, 1.41). The study by Rahimi et al. contributed the least weight while that of Li-Xin et al. 2013 had the largestcontribution to the overall weight^46,49^. In addition, the heterogeneity has a Chi^2^ with a p-value of 0.0001 and I^2^= 83%. The p-value of the overall/summary effect is 0.09 which is statistically non-significant.

Figure 5.0 shows the result of the meta-analyses after a sensitivity analysis was carried out to determine the cause of high heterogeneity where the effect of curcumin on fasting blood glucose was the outcome measured. The study by Rahimi et al. 2016 was identified and removed from the analysis on the basis of dose variability. The test for heterogeneity showed Chi^2^ with p = 0.32 and I^2^=15% indicating low heterogeneity. The pooled effect estimate shows a total 95% confidence interval of -1.84(-4.92, 1.24) which favours the curcumin group. The total sample size was 279 and an overall p-value of 0.24 showed a lack of statistical significance^46^.

The second forest plot in figure 6.0 shows the result for the effect of curcumin on glycated haemoglobin concentration (HbA1c). All the point estimates for individual studies fell to the left of the line of no effect in favour of curcumin. All the studies also had narrow confidence intervals indicating a more precise result. The study by Selvi et al. contributed the greatest weight while the study by Hodaei et al. 2019 contributed to the lowest weight. The overall test effect fell to the left of the line of no effect with a p-value of 0.05 which shows statistical significance. The overall mean difference/95% confidence interval was -0.58 (-1.15, -0.01). Heterogeneity showed a Chi^2^ with a p-value of 0.02 and I^2 =^ 66% which indicates high variability across the included studies^43,44^.

Figure 7.0 shows the result where the effect of curcumin on HbA1c was the outcome measured following sensitivity analysis. The study by Rahimi et al.2016 was removed due to a very low dose of curcumin (80mg) compared to the other studies whose dose ranged from 300mg/day to 2100mg/day. The test for heterogeneity showed a Chi^2^ with p = 0.41 and I^2^= 0% showing a complete lack of heterogeneity. The test for overall effect shows a lack of statistical significance with a p-value of 0.13. The pooled estimate following the sensitivity analysis showed a total 95% confidence interval of -0.21(-0.55, 0.07) with the diamond still falling to the left of the line of no significance in favour of curcumin supplemented group^46^.

## Results from funnel plots

Figures 8.0 and 9.0 below shows the funnel plots for fasting blood glucose before and after sensitivity analysis was carried out respectively. Figures 10.0 and 11.0 shows the funnel plots for HbA1c before and after the conduct of a sensitivity analysis respectively. This was conducted to demonstrate the possibility of publication bias in the included studies. Results from all the funnel plots are from studies conducted using the random effect model. The circles represent individual studies while the dotted vertical line represents the total average in which 95% of studies lies. The standard error (or a measure of precision) on the Y-axis is plotted against the treatment effect of the individual studies on the X-axis. Smaller studies with less precision lie towards the bottom of the graph while larger studies with greater precision sit at the top of the graph. Results show a skewed funnel or asymmetry of the plot indicative of a potential risk of bias. The outlier in figure 6 shows the study by Rahimi et al. 2016. Its position away from the average is indicative of less precision of the study which can be due to smaller sample size.

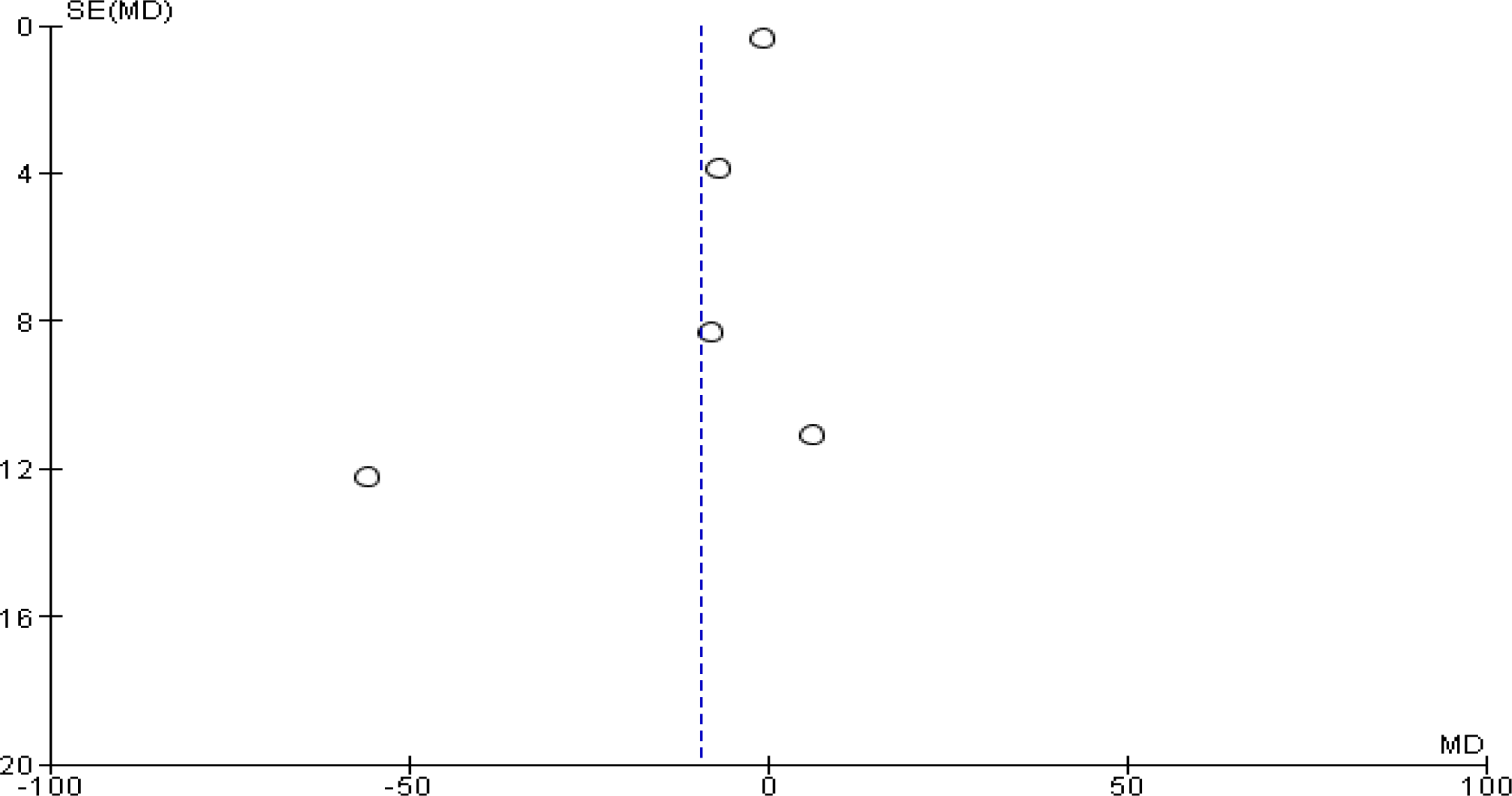
Funnel plot of comparison on FBG before sensitivity analysis.

**Figure 9.0:**
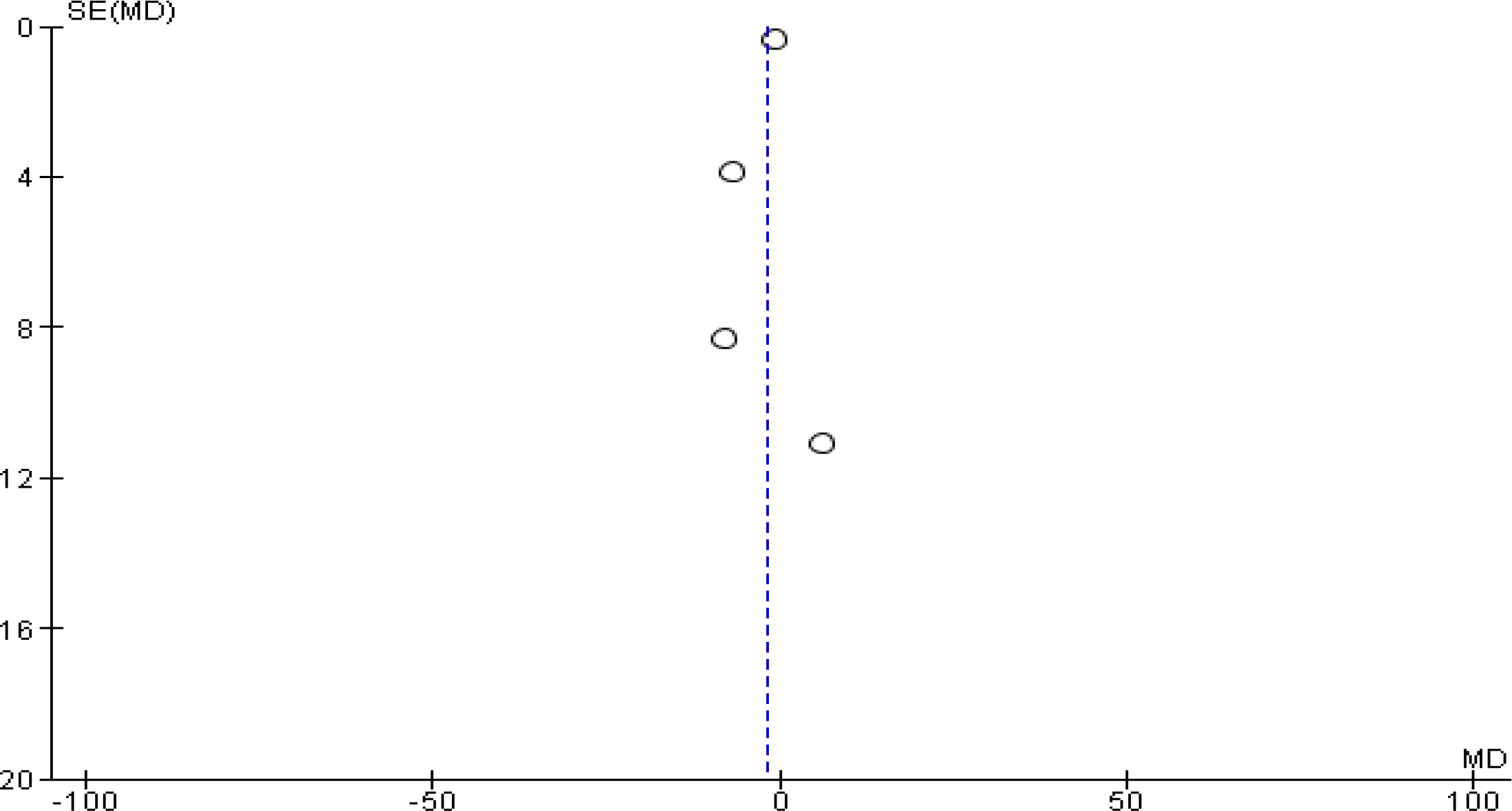
Funnel plot of comparison on FBG after sensitivity analysis

**Figure 10.0:**
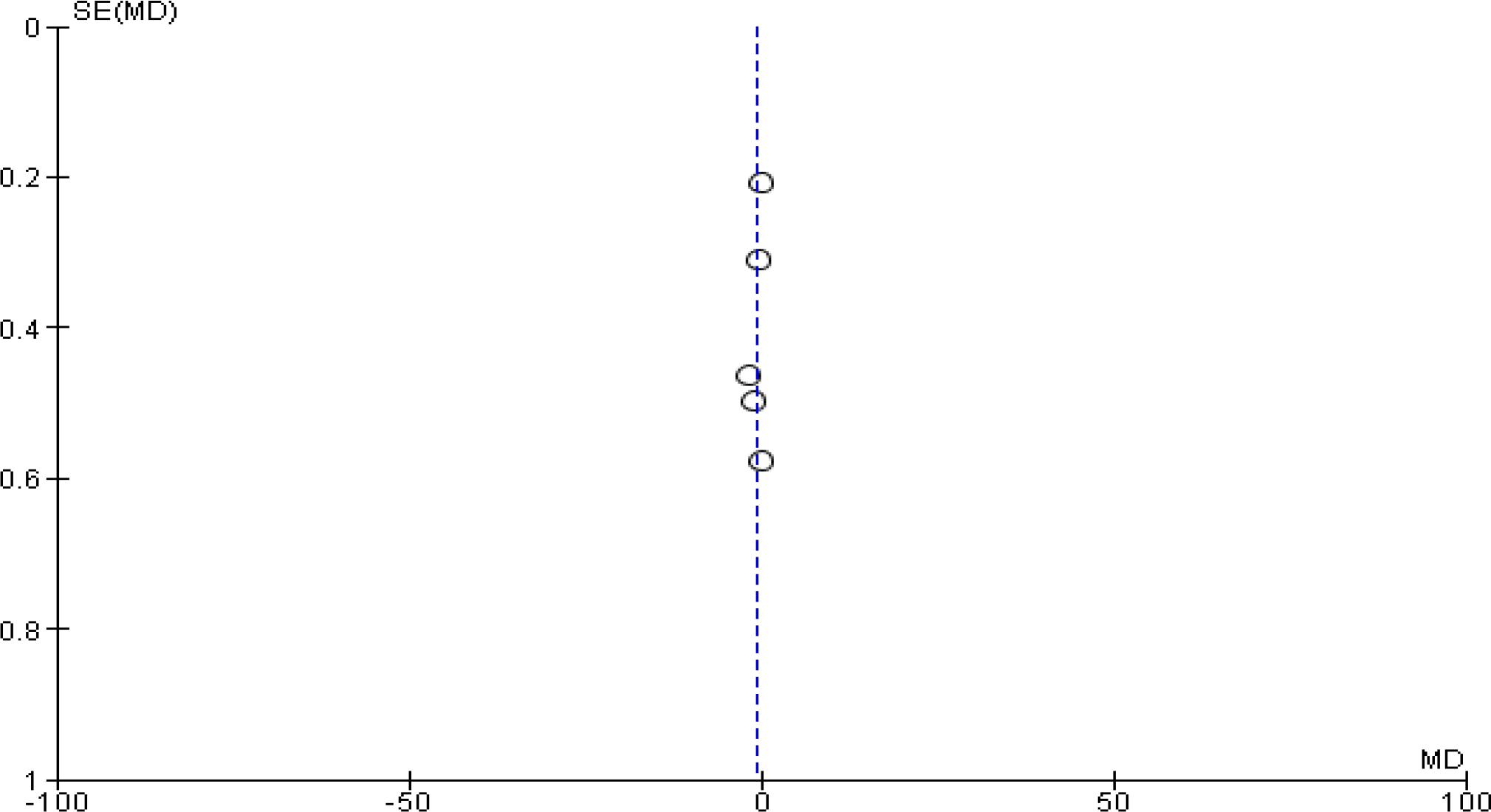
Funnel plot of comparison on HbA1c before sensitivity analysis

**Figure 11.0:**
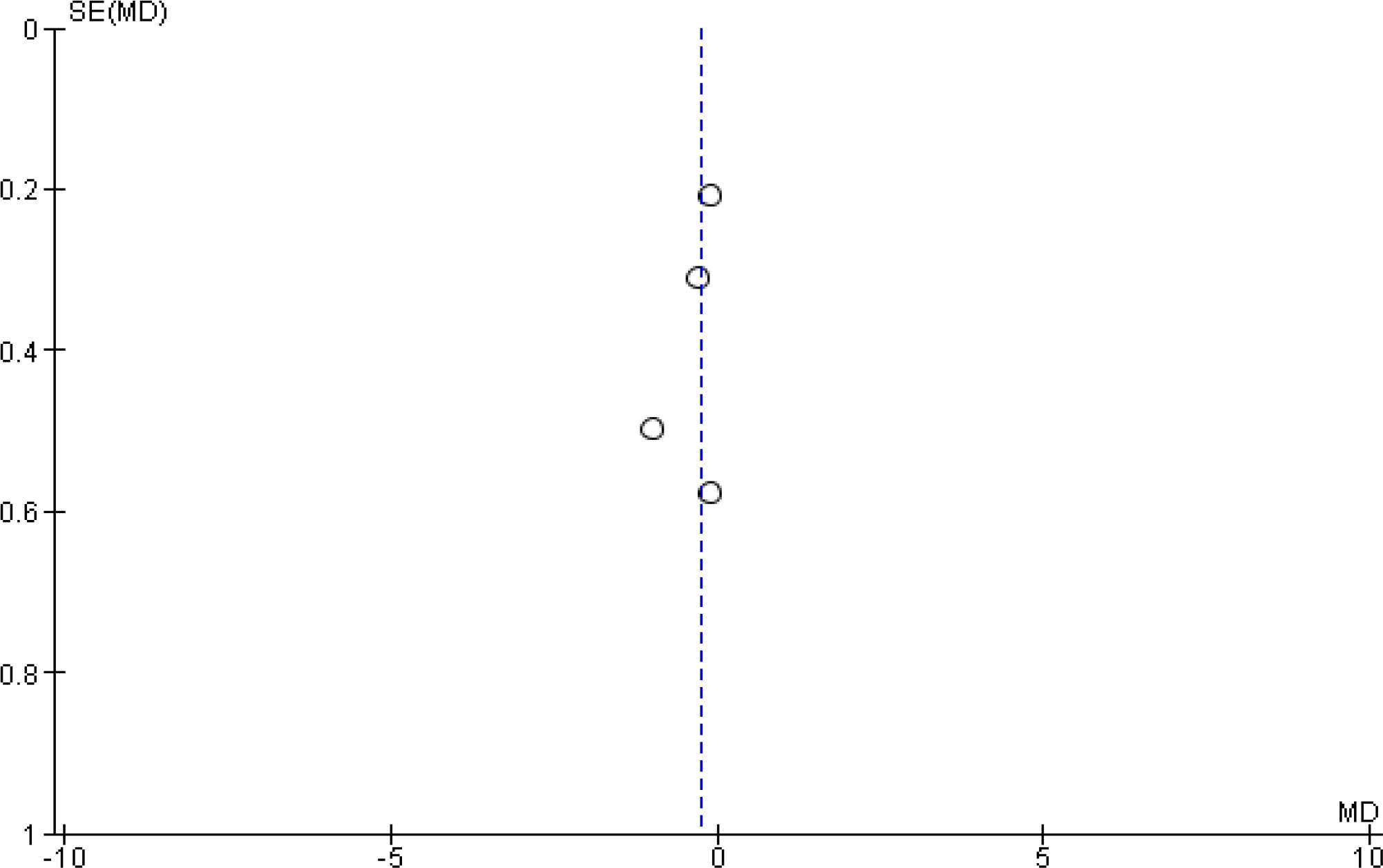
Funnel plot of comparison on HbA1c after sensitivity analysis

## 4.0 DISCUSSION

This study was aimed at using a systematic review methodology to determine the effect of curcumin (turmeric) on fasting blood glucose and glycated haemoglobin in patients with type 2 diabetes mellitus. Evidence shows that there is a causal relationship between oxidative stress, inflammation, hyperglycaemia and other glycaemic indices in the pathogenesis and progression of T2DM^49^. Since the current chemical hypoglycaemic agents have limited efficacy in eliminating free radicals and inflammatory processes, there is a growing interest to usenatural polyphenols such as curcumin from turmeric as an adjunct intervention in diabetes management. This is because curcumin hasdemonstrated anti-oxidative and antiinflammatory effects.

The choice of FBG and HbA1c as the outcomes to be measured was informed by their diagnostic importance and reliability in determining the efficacy/effectiveness of interventions in the management of diabetes^40^.

Given that the few studies conducted on human participants come with a lot of mix evidence, conducting a systematic review and meta-analyses using RCTs was deemed appropriate to establish stronger evidence on the effectiveness of curcumin that could help inform a clinical decision, inspire further studies and guide policymaking.

From the result of the meta-analyses on the effect of curcumin on fasting blood glucose, all the included studies and the pooled effect estimate except the study by Hodaei et al. 2019 favoured curcumin^44^. This shows that the use of curcumin as an adjunct therapy significantly reduced fasting blood glucose. This result agrees with a previous systematic review conducted by Poolsup et al. involving prediabetics and diabetics, this study also showed that curcumin significantly reduced FBG^37^. The antihyperglycemic effect of curcumin has shown great similarity with a thiazolidine-dione group of oral hypoglycaemic drugs and even much more is the antioxidative and anti-inflammatory effects^50^. It has the ability to activate PPAR-□ which helps to enhance insulin secretion thus, a significant effect on glycaemic control^51^. The study by Jiménez-Osorio, Monroy, & Alavez, 2016 also showed that curcumin administered to diabetic patients had the ability to induce the expression of glucose transporter-4 (GLUT-4) independently which in turn enhanced glucose uptake from the blood hence, the control of hyperglycaemia^52^.

The greatest reduction in FBG was observed in the study by Rahimi et al. (2016) as observed from the point estimate shown on the forest plot in figure 4.0 and figure 4.1. This effect is observed despite that the participant in this study received the lowest dose of curcumin (80mg administered once daily)^46^. This could possibly be associated with the combination of curcumin and Nano-micelles to enhance its bioavailability. This result further suggests that curcumin delivered at higher doses of more than 2000mg produced a lesser effect compared to when nanocurcumin was used even at a much lower dose. This is possibly due to its poor bioavailability. Studies have shown that curcumin has poor bioavailability when taken orally because of its susceptibility to temperature changes, stomach acidity, enzymatic actions, and poor solubility^53^. This may be responsible for the non-significant result observed in some previous studies where only curcumin/turmeric powder wasused. Research in medical Nanotechnology proved that a combination of curcumin with Nano-micelles (nanocurcumin) increased curcumin’s bioavailability when compared to turmeric powder or curcumin alone^46^. Other delivery methods that have been studied and reported to increase the bioavailability of curcumin include liposomal curcumin, microemulsions and polymeric implantable devices^54^.

Although there was a distinct clinical significance in the reduction of FBG as observed from the pooled effect estimate, there was no statistical difference between the curcumin supplemented group and the control group. This is also demonstrated where the overall confidence interval slightly crossed the line of no significance. This does not necessarily mean a lack of effectiveness but could mean that the total sample size was too small and further studies may need to be carried out in a larger sample size to further re-affirm this evidence. More so, the wider confidence interval in this particular study represents a less precise result and a smaller sample size which may also potentiate the effect of curcumin. Button et al. (2013) observed that studies with smaller sample size are likely to show increased effect in the experimental group while findings from studies with larger sample size are more likely to represent the true effect. Therefore, with larger sample size and appropriate delivery method for pure curcumin to bypass bioavailability issues, the overall effect of curcumin as an adjunct therapy in diabetes will be promising as indicated in the overall results^55^.

Assessment of the effect of curcumin on HbA1c was also carried out. The resultdisplayed in figure 5.0 and figure 5.1 above showed that the point estimates for all the included studies were towards the left of the line of no effect indicating that curcumin decreases HbA1c in type 2 diabetic patients. The pooled effect also demonstrated a reduction in HbA1c in favour of the curcumin supplemented group. This finding is very important because studies have shown that higher levels of HbA1c indicate a higher risk of developing diabetic complications and pancreatic tissue damage^56^.The result showed that participants in the curcumin group were more likely to have a higher reduction in the HbA1c concentration than those in the control group who received the usual chemical hypoglycaemic drugs alone. This also means that patients administered curcumin in addition to their usual diabetic drugs will most likely prevent the progression of T2DM and the development of diabetic complications than their counterparts receiving only the usual diabetic drugs. Curcumin’s preventive and reparative functions are linked with its an-oxidative and anti-inflammatory effects in the insulin-signalling pathway and glucose metabolism^57^. These properties are largely linked with the maintenance of vascular integrity or the repair of damaged vascular endothelia which is a major step in the development of diabetic complications.

It must be noted that HbA1c measures the percentage of glucose bound to haemoglobin which predicts glycaemic control over a 90-120 days period (the average lifespan of the red blood cells). However, all the studies had a treatment duration of ≤ 90 days which may underestimate the treatment effect of curcumin. Therefore, studies with treatment duration ≥ 3 months would have been more suitable. Studies with shorter treatment duration like the study by Selvi et al. have the likelihood of underestimating the effect of curcumin on HbA1c.

Using human red blood cells model exposed to high glucose to mimic diabetes *in vitro*, curcumin prevented the glycosylation of protein and increased utilisation of glucose by the cells^58^. This suggests that at*in vivo* conditions, curcumin is able to decrease HbA1c concentration thus, the control of hyperglycaemia and the prevention of cardiovascular complications in type 2 diabetic patients. Since most chronic diseases including diabetes involve multiple pathways, curcumin is advocated as an adjunctive intervention because evidence shows its multitargeting effect on different molecular and metabolic pathways in the body^59^.

The test for heterogeneity presented a p-value of 0.0001 and I^2^ value of 83% which are both ≤ 0.1 and ≥ 50% respectively thus, a random effect model was used to analyse data. Boland et al. (2017) suggested that a random effect model is more appropriate where heterogeneity is suspected to be high. This is because using a fixed-effect model, in this case, will invalidate the “assumption that there is only one underlying true effect measured across all the trials”^43^. The limitation of this model, however, is that smaller studies tend to produce more effect in a random effect model^60^. Although the quality assessments showed that all the included studies were high-quality research, the result of heterogeneity demonstrated a high degree of variation across the different studies. These variations are suggested to be attributable to differences in dose (80mg to 2100mg), sample size and duration of treatment (from 4 weeks to 12 weeks across studies). There was also variation in the form of delivering the intervention i.e. either as turmeric powder, pure curcumin or nanocurcumin which can also influence the treatment effects observed in the individual studies and the metaanalysis.Evidence hasshown that only 2%-6% of turmeric powder will contain curcumin (the most important component of turmeric for diabetes management) therefore, studies with turmeric powder tends to show a lower effect when compared with studies where pure curcumin is extracted and used as the intervention^59^. Enhancing the bioavailability of curcumin only means that lower doses can be used to achieve greater therapeutic effects.

Although the inclusion criteria involved diabetic patients greater than 18 years of age, this wider age range between 18-75 years means that there could be variations in the metabolism of curcumin and response to treatment. Shalini (2019) explained that infants and elderly patients usually have a lower response to drugs when compared to middle-age patients despite the effectiveness of such intervention. Therefore, age variation can also contribute to high heterogeneity in a study^61^.

It has been suggested that when the Chi^2^ and I^2^ (i.e. the degree of variation across studies is very high), a sensitivity analysis should be carried out to find out the cause of the high heterogeneity^43^. In this study, strict criteria were initially considered on a minimum dose of 300mg and delivering form involving either turmeric powder or curcumin. However, due to a limited number of studies on the intervention and outcome of interest, the study by Rahimi et al. (2019) was included despite using a dose of 80mg/day and Nano-micelles to enhance bioavailability. Therefore, a sensitivity analysis was conducted to exclude the study by Rahimi to test the robustness of the result and confirm the pooled effect estimate from the meta-analysis as explained by Boland, Cherry & Dickson (2017).

There was high heterogeneity observed in both outcome measures (fasting blood glucose and HbA1c which demonstrated a high degree of the variability across the five studies included in the meta-analyses. Further investigations were carried by removing studies on the basis of duration, sample size, age, and form of delivering curcumin to know the cause of high heterogeneity. All results still showed high heterogeneity (i.e. I^2 >^50%).

Following a sensitivity analysis involving the removal of the study by Rahimi et al. (2019) on the basis of dose variation, the heterogeneity was very low (I^2^ = 15%) for analysis on fasting blood glucose and there was no variability (I^2^ = 0%) observed in the analysis of HbA1c. This result confirmed that the study by Rahimi et al. (2019) in which nanocurcumin was used and given at a relatively low dose contributed greatly to the high heterogeneity observed initially^46^. However, the low or lack of variability observed after the sensitivity analysis showed higher p values indicative of smaller sample size which has the potential of positively or negatively influencing the overall effect of curcumin on both FBG and HbA1c.

It must also be noted that an I^2^ of 0% does not necessarily mean the absence of variability. It could simply mean that the number of studies included in the meta-analysis was very small which can underestimate the variability. von Hippel (2015) conducted a study on metaanalyses submitted in the Cochrane library and explained that substantial bias in I^2^ is observed in meta-analyses with small studies i.e. the number of included studies less than 7. von Hippel showed that reviews with ≤7 included studies have the potential of underestimating the heterogeneity by an average of 28%.Therefore, the more the number of

included studies, the better the true estimate of I-squared statistics.However, the total effect estimate from the sensitivity analysis still confirms that type 2 diabetic patients in the curcumin supplemented group were more likely to have a significant reduction in fasting blood glucose and HbA1c than those treated with the conventional diabetic medication (i.e. the control group) irrespective of dose or form of delivering of curcumin^62^.

The reviewer of this study also observed that studies that use the pure curcumin had a significant reduction in fasting blood glucose and HbA1c when compared to studies by Selvi et al. (2015) where turmeric powder was used^43^. A similar meta-analysis conducted by Poolsup et al. (2019) showed that pure curcumin had a greater effect on fasting blood glucose and HbA1c than turmeric in both diabetic and non-diabetic patients. Similarly, studies with longer duration i.e. ≥ 8 weeks produced a more significant reduction in FBG and HbA1c when compared with the study by Selvi et al. (2015) with only 4 weeks of treatment which showed a lesser effect^43^. Adab et al. (2013) suggested that a minimum of 8 weeks of treatment with curcumin is recommended to produce significant benefits in glycaemic control^47^.

### Publication bias

In this study, although individual studies were of higher quality for systematic review and meta-analyses as demonstrated in the quality assessment, there were indications of publication bias given the skewness or asymmetry of the funnel plots. High-quality studies do not necessarily translate to an absence of publication bias since this kind of bias can result from hypothesis testing^63^. For instance, studies with statistically significant results are more likely to be published in journals than studies with null results. However, Sterne, Gavaghan, & Egger (2000) explained that the skewness of a funnel plot can result from other reporting bias other than publication bias. These include location bias, language bias, non-response bias and citation bias, or bias resulting from the systematic difference in the included studies or methodological design of the primary studies. Also, the overestimation of treatment effect in smaller studies can result in asymmetry of the funnel plot^63^.

In the current study, the skewness or asymmetry of the funnel could be attributed to the absence of larger studies or failure to publish studies with a null result. Lack of access or the omission of unpublished studies from a systematic review can lead to bias in the effect estimate of the intervention^64^. Secondly, this could be linked with the systematic variability among included studies. Sterne et al. (2000) reiterated that the asymmetry of the funnel plot can result from high heterogeneity across the included studies^63^.

### Limitations and Recommendations

There are limitations observed in the current study. One such limitation is that all the included studies in this analysis are of Asian origin. This is possibly due to the popularity of curcumin/turmeric used for both culinary and medicinal purposes in the continent. However, it is recommended that other studies are conducted in other locations to prevent reporting bias such as location bias.

In both individual studies and the meta-analyses, there were indications of small sample size creating a lack of statistical significance.Therefore, more studies with larger sample size are recommended in order to increase the precision and strength of evidence as well as the applicability of the result to the general population.

Studies with pure curcumin extract were initially considered but because of the very limited number of studies,the inclusion criteria were expanded to include studies where curcumin in the form of turmeric powder and nanocurcumin were given to the treatment group. This contributed to the dose variability across studies as demonstrated by the sensitivity analysis.

It will be more appropriate to conduct studies with specific delivery form (e.g. pure curcumin extract alone or Nano-curcuminalone to increase the validity of the results.

Lastly, there were two quality studies that were not included because of inaccessibility to the articles and inability to reach out to the original authors, which could have increased the overall sample size adding to the statistical power of the pooled effect estimate.

## Conclusion

This study showed that curcumin, a polyphenolic compound extracted from turmeric is effective in reducing fasting blood glucose and the levels of glycated haemoglobin in patients with T2DM. This is majorly due to its antioxidative and anti-inflammatory properties as well as its ability to influence numerous metabolic pathways in the body.

The clinical implication of this result is that the clinical use of curcumin as an adjunct therapy has beneficial roles over the common chemical hypoglycemic drugs in the management of hyperglycaemia, T2DM, and the prevention of diabetic complications. This study also reveals that the clinical use of curcumin may be hampered by its poor bioavailability thus, the combination of curcumin with Nano-micelles or other hydrophilic agents will help bypass these challenges and maximize its therapeutic potential in diabetics. Treatment for at least 8 weeks also tends to produce more effect than treatment of shorter durations.

The paucity of scientific knowledge on the effect of curcumin in patients with T2DM and the limitations to this systematic review implies that more research needs to be conducted with special consideration to issues around the effective dose, study settings, therapeutic duration, best delivery form of pure curcumin extract, and larger sample size in order to firmly establish its effectiveness and applicability in the larger population. However, curcumin may prove to be safer, cheaper, relatively available and a very potent supportive intervention to help curb the global health crisis posed by type 2 diabetes mellitus.

## Data Availability

All data produced in the present study are available upon reasonable request to the authors

## Ethics Approval and consent to Participate

Not Applicable **Consent to Publish** Not applicable

## Availability of Data and Materials

The Data set from the study are available to the corresponding author upon request.

## Competing Interests

Authors have declared that they have no competing interests

## Funding

No funds were received for this study

## Acknowledgements

Not Applicable

## Notes

### Competing Interest Statement

The authors have declared no competing interest.

### Funding Statement

This study did not receive any funding

